# Meta-Analysis of Adenoviral p53 Gene Therapy Clinical Trials in Recurrent Head and Neck Squamous Cell Carcinoma

**DOI:** 10.1101/2021.01.06.20248743

**Authors:** Robert E. Sobol, Kerstin B. Menander, Sunil Chada, Dora Wiederhold, Beatha Sellman, Max Talbott, John J. Nemunaitis

## Abstract

**Background:** We conducted a meta-analysis of previous adenoviral p53 (Ad-p53) treatment data in recurrent head and neck squamous cell carcinoma (HNSCC) patients to identify optimal Ad-p53 treatment methods for future clinical trials.

**Methods:** The meta-analysis involved recurrent HNSCC patients treated with Ad-p53 for whom p53 genotyping and immunohistochemistry tumor biomarker studies had been performed (n = 70). Ad-p53 tumor treatment responses defined by RECIST 1.1 criteria were correlated with Ad-p53 dose and tumor p53 biomarkers. Gene expression profiles induced by Ad-p53 treatment were evaluated using the Nanostring IO 360 panel.

**Results:** Ad-p53 dose based upon the injected tumor volume had a critical effect on tumor responses. All responders had received Ad-p53 doses greater than 7 × 10^10^ viral particles/cm^3^ of tumor volume. There was a statistically significant difference in tumor responses between patients treated with greater than 7 × 10^10^ viral particles/cm^3^ compared to patients treated at lower Ad-p53 doses (Tumor Response 31% (9/29) for Ad-p53 > 7 × 10^10^ viral particles/cm^3^ versus 0% (0/25) for Ad-p53 < 7 × 10^10^ viral particles/cm^3^; p = 0.0023). All responders were found to have favorable p53 biomarker profiles defined by less than 20% p53 positive tumor cells by immunohistochemistry (IHC), wild type p53 gene sequence or p53 deletions, truncations, or frame-shift mutations without functional p53 tetramerization domains. Preliminary gene expression profiling results revealed that Ad-p53 treatment increased Type I Interferon signaling, decreased TGF-beta and beta-catenin signaling resulting in an increased CD8+ T cell signature which are associated with increased responses to immune checkpoint blockade.

**Conclusions:** Our findings have important implications for future p53 targeted cancer treatments and identify fundamental principles to guide Ad-p53 gene therapy. We discovered that previous Ad-p53 clinical trials were negatively impacted by the inclusion of patients with unfavorable p53 biomarker profiles and by under dosing of Ad-p53 treatment. Future Ad-p53 clinical trials should have favorable p53 biomarker profiles inclusion criteria and Ad-p53 dosing above 7 × 10^10^ viral particles/cm^3^ of injected tumor volume. Preliminary gene expression profiling identified p53 mechanisms of action associated with responses to immune checkpoint blockade supporting evaluation of Ad-p53 in combination with immune checkpoint inhibitors.

## Background

Head and neck cancers represent the sixth most common cancer world-wide with approximately 630,000 patients and 350,000 deaths annually(1,2). Greater than 90% of the cases are squamous cell carcinomas arising from the oral cavity, oropharynx and larynx. Numerous investigations regarding the cause and progression of human cancer have identified the loss of p53 tumor suppressor function as a major pathogenetic factor in most tumor types including HNSCC(3). TP53 is the prototypic tumor suppressor that mediates a wide range of functions including cell cycle arrest, DNA damage repair, cellular senescence, apoptosis, and epithelial-mesenchymal transition (EMT) (4). A replication defective adenoviral vector containing the wild type p53 gene (Ad-p53) and its necessary expression cassette was constructed and has been employed in multiple clinical trials(5) (6)(7)(8)

In this investigation, we performed a meta-analysis of previous Ad-p53 monotherapy clinical trials in recurrent HNSCC correlating tumor response with biomarkers, dosing and administration methods. In addition to the previously reported p53 efficacy biomarkers(7), we identified Ad-p53 dosing parameters based upon the number of viral particles per mm3 of tumor as a critical predictor of efficacy. We also report the initial results of nanostring gene expression profile changes associated with Ad-p53 treatment in a HNSCC patient with a dramatic response to combined Ad-p53 and immune checkpoint inhibitor blockade.

## Methods

### Meta-Analysis of Ad-p53 Clinical Trials in Recurrent HNSCC

A replication defective adenoviral vector containing the normal p53 gene (Ad-p53) and its expression cassette using a CMV promoter was constructed as described (5). In previous Ad-p53 gene therapy clinical trials in recurrent HNSCC, Ad-p53 was administered in treatment schedules of three times per week intratumorally either as three consecutive daily Ad-p53 treatments during the first week or every other day for the first 2 weeks of each monthly treatment cycle. In these studies, a uniform dose of 2 x 10^12^ viral particles per treatment was administered intra-tumorally(7). The dose was divided between the patients’ tumors at the investigators’ discretion which lead to different tumors receiving different Ad-p53 doses by tumor volume. In this meta-analysis, we evaluated the dose of Ad-p53 administered per tumor volume using the bi-dimensional tumor diameters of length (longer diameter) and width (shorter diameter) in the formula Tumor Volume = (0.5)(L)(W^2^). Tumor response was assessed by RECIST 1.1 criteria(9). Earlier Ad-p53 gene therapy studies identified a predictive p53 biomarker profile of Ad-p53 therapeutic efficacy(7). This favorable profile (wild type p53 gene sequence or less than 20% p53 protein positive tumor cells by immunohistochemistry) predicted treatment efficacy and identified HNSCC patients more likely to benefit from Ad-p53 therapy with increased tumor responses and improved survival(7); (8)

To identify optimal Ad-p53 treatment doses for future trials, we conducted a meta-analysis of the previous Ad-p53 treatment data in recurrent HNSCC patients(7,8). The meta-analysis involved recurrent HNSCC patients where tumor p53 genotyping and immunohistochemistry biomarker studies had been performed (n = 70). Ad-p53 was administered in treatment schedules of three times per week intratumorally either as three consecutive daily Ad-p53 treatments during the first week or every other day for the first 2 weeks of each monthly treatment cycle. The demographic and baseline characteristics of these patients have been described previously(7).

### Biomarker Studies

#### p53 and Other Biomarker Profiles

TP53 protein expression by immunohistochemistry and p53 genotyping were performed as described previously (7); (8) A similar immunohistochemistry assay was employed to detect PD-L1 expression and was performed by Cancer Genetics Incorporated, Rutherford, NJ. Foundation One CDx was utilized to determine microsatellite instability (MSI), tumor mutational burden (TMB) and other biomarkers (Foundation One, Cambridge, MA).

#### Nanostring Transcriptome Gene Expression Analyses of Ad-p53 Treatment

We describe in this report, the initial transcriptome results of gene expression profiles induced by Ad-p53 treatment performed as part of a new Ad-p53 recurrent HNSCC clinical trial in combination anti-PD-1 therapy NCT03544723 (https://clinicaltrials.gov/ct2/show/NCT03544723). RNA was isolated from pre- and post-treatment samples and compared using Nanostring IO 360 gene expression panel (Nanostring Technologies Seattle WA). This panel tests expression of 770 genes involved in neoplasm pathology, tumor microenvironment and cancer immune responses. Samples were processed and analyzed as described (Nanostring PanCancer <u>IO360 Best Practices Guide</u>).

## Results

### Meta-Analysis of Ad-p53 Monotherapy Data in Recurrent HNSCC

To identify optimal Ad-p53 treatment doses for future trials, we conducted a meta-analysis of the previous Ad-p53 treatment data in recurrent HNSCC patients. The meta-analysis involved recurrent HNSCC patients where p53 biomarker studies had been performed (n = 70). In the meta-analysis, all responders were found to have favorable p53 biomarker profiles (wild type p53 gene sequence or less than 20% p53 protein positive tumor cells by immunohistochemistry). We therefore conducted the meta-analysis of the Ad-p53 treatment data in recurrent HNSCC patients with favorable Ad-p53 biomarker profiles (n = 54). This approach eliminated patients with unfavorable p53 biomarker profiles (n = 16) defined by high level expression of dominant negative mutated p53, which form mixed p53 tetramers that could inhibit the activity of wild-type p53(7). The demographic and baseline characteristics of these patients have been described previously and there were no statistically significant differences between patients with favorable and unfavorable p53 biomarker profiles(7).

In these prior Ad-p53 clinical trials, a total dose of 2 x 10^12^ viral particles per treatment was divided between the patients’ tumors at the investigators’ discretion, which lead to different tumors receiving different Ad-p53 doses by tumor volume. In this meta-analysis, we evaluated the dose of Ad-p53 administered per injected tumor volume using the formula Tumor Volume = (0.5)(L)(W^2^) where L= Length of longer diameter and W=width of shorter diameter. As shown in Figure 1, Ad-p53 dose based upon the injected tumor volume had a critical effect on tumor response. All Ad-p53 treated target lesion responders, by RECIST 1.1 criteria, had received Ad-p53 doses greater than 7 × 10^10^ viral particles/cm^3^ tumor.

**Figure 1.**
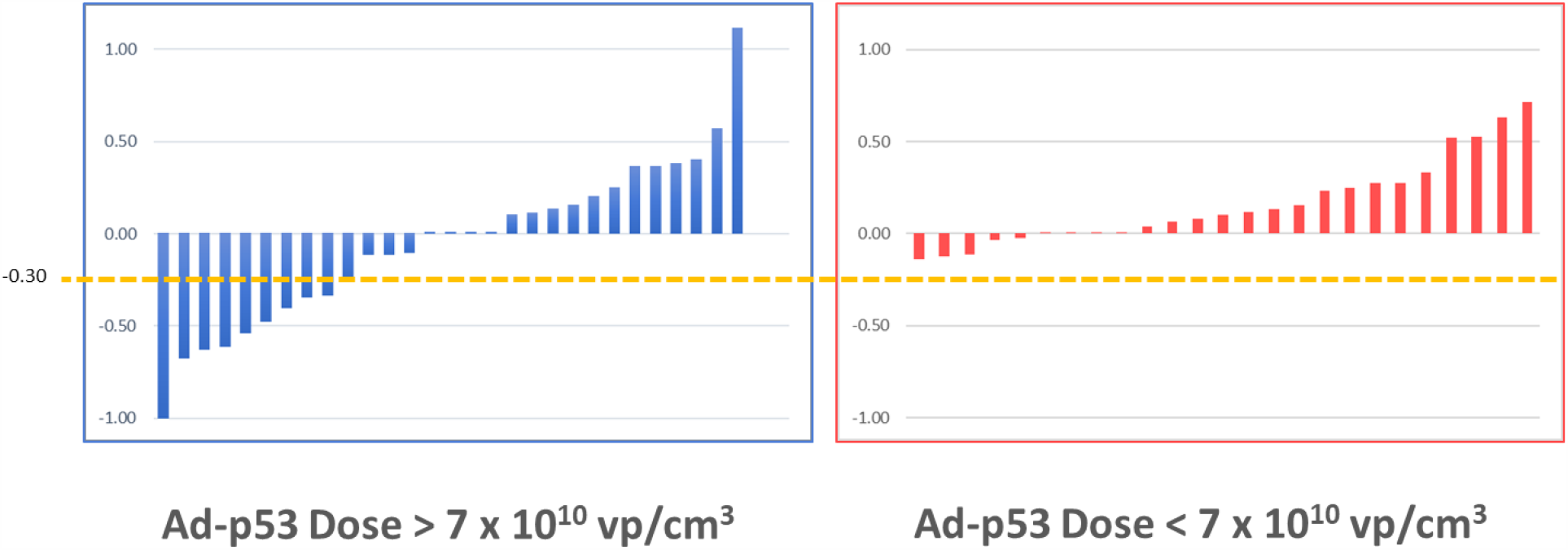
Waterfall plot of percent change in tumor size of target lesions following treatment with Ad-p53. All tumor lesion responses by RECIST 1.1 (reduction of at least 30%) occurred in patients Ad-p53 doses greater than 7 x 10^10^ viral particles/cm^3^ of tumor volume.

In the group receiving greater than 7 × 10^10^ viral particles/cm^3^ tumor, 31% (9/29 patients) were responders compared to 0% (0/25 patients) treated with less than 7 × 10^10^ viral particles/cm^3^ tumor. This difference was statistically significant p = 0.0023 by Fisher’s exact test.

#### Gene expression profiles induced by Ad-p53 treatment

A more detailed examination of the Ad-p53 response data revealed that the majority of responders (7/9 patients) had received doses of Ad-p53 near or exceeding 1 x 10^11^ vp/cm^3^ (range 7.81 to 333.2 × 10^10^ vp/cm^3^). Hence, based upon this data, an Ad-p53 dose of 1 x 10^11^ vp/cm^3^ of tumor volume was chosen for a new trial combining Ad-p53 with anti-PD-1 therapy NCT03544723 (https://clinicaltrials.gov/ct2/show/NCT03544723. Anti-PD-1 therapy has become the standard of care for patients with recurrent HNSCC who have progressed after platinum chemotherapy(10). Ad-p53 was administered by intra-tumoral injections on days 1, 2, and 3 of each cycle and Nivolumab (480 mg) was administered as an intravenous infusion every 28 days starting on Day 5 of the first study cycle.

The pre and post Ad-53 treatment transcriptome gene expression profiling results reported here are for the first patient treated in this Phase 2 clinical study. The patient is a male in his sixth decade who was originally diagnosed with early stage, T1N0M0, head and neck squamous cell cancer (HNSCC) of the right tonsil following a bilateral tonsillectomy. The pathology showed moderately differentiated squamous cell carcinoma, with diffusely positive P16 staining, consistent with a human papilloma virus (HPV) associated carcinoma. The tumor was reported to focally involve a margin, however, he declined adjuvant radiotherapy. He was found to have recurrence two years later with nodal metastases and he was treated with a chemotherapy regimen consisting of carboplatin, doxetaxel, and ifosfamide. He had evidence of further progression after this treatment, and then pursued enrollment in clinical trials.

FoundationOne CDx analyses performed in at study entry indicated the pre-treatment lesion was p53 wild type and had mutations in NF1, PI3KCA, EZH2, NFE212 and MLL2 genes. There were no mutations in mismatch repair genes (MMR) or microsatellite gene markers, indicating his status as mismatch repair proficient (pMMR) and microsatellite stable. The Tumor Mutational Burden was 13 Muts/Mb, reflecting an Intermediate/Low status(11).

p53 protein levels were also assessed by immunohistochemistry using DO-7 antibody (Ventana), and the pre-treatment sample was 15% positive for wild type p53 protein and using the 22C3 antibody to detect PDL-1 showed 5% positive staining.

To assess the gene expression profiles induced by Ad-p53 treatment, pre and post treatment biopsies after two cycles of therapy were obtained and compared using Nanostring IO 360 gene expression panel. Figure 2 shows the dramatic clinical response in the lesion evaluated for changes in gene expression.

**Figure 2.**
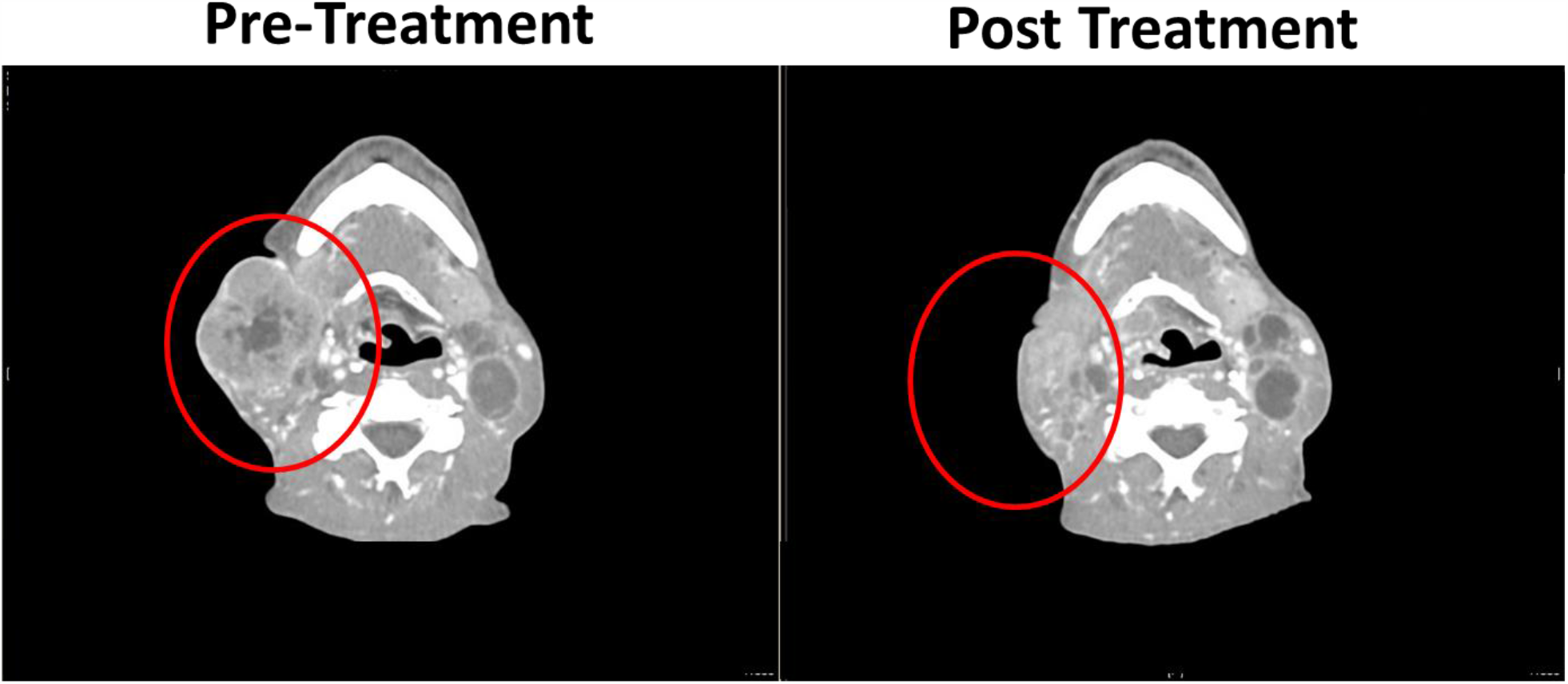
Pre and post Ad-p53 treatment computerized tomography scans of responding tumor lesion utilized for RNA gene expression transcriptome analyses by nanostring IO 360 panel.

RNA was isolated from the pre- and post-treatment samples and compared using the Nanostring IO 360 gene expression panel. This panel tests expression of 770 genes and evaluates genes involved in the complex interplay between the tumor, its microenvironment and the anti-tumor immune response. The panel incorporates 47 biological signatures and includes the 18-gene Tumor Inflammation Signature (12)associated with response to PD-1/PD-L1 inhibitors pathway blockade. The pre-treatment lesion had a Tumor Inflammation Score (TIS) of 6.2 and post-treatment lesion was 7.1. As shown in Figure 3, the key gene expression profiling results revealed that Ad-p53 treatment increased the Tumor Inflammation Signature and Type I Interferon signaling, decreased TGF-beta and beta-catenin signaling resulting in an increased CD8^+^ T cell signature which is associated with increased clinical responses to immune checkpoint inhibitor therapy(12).

**Figure 3.**
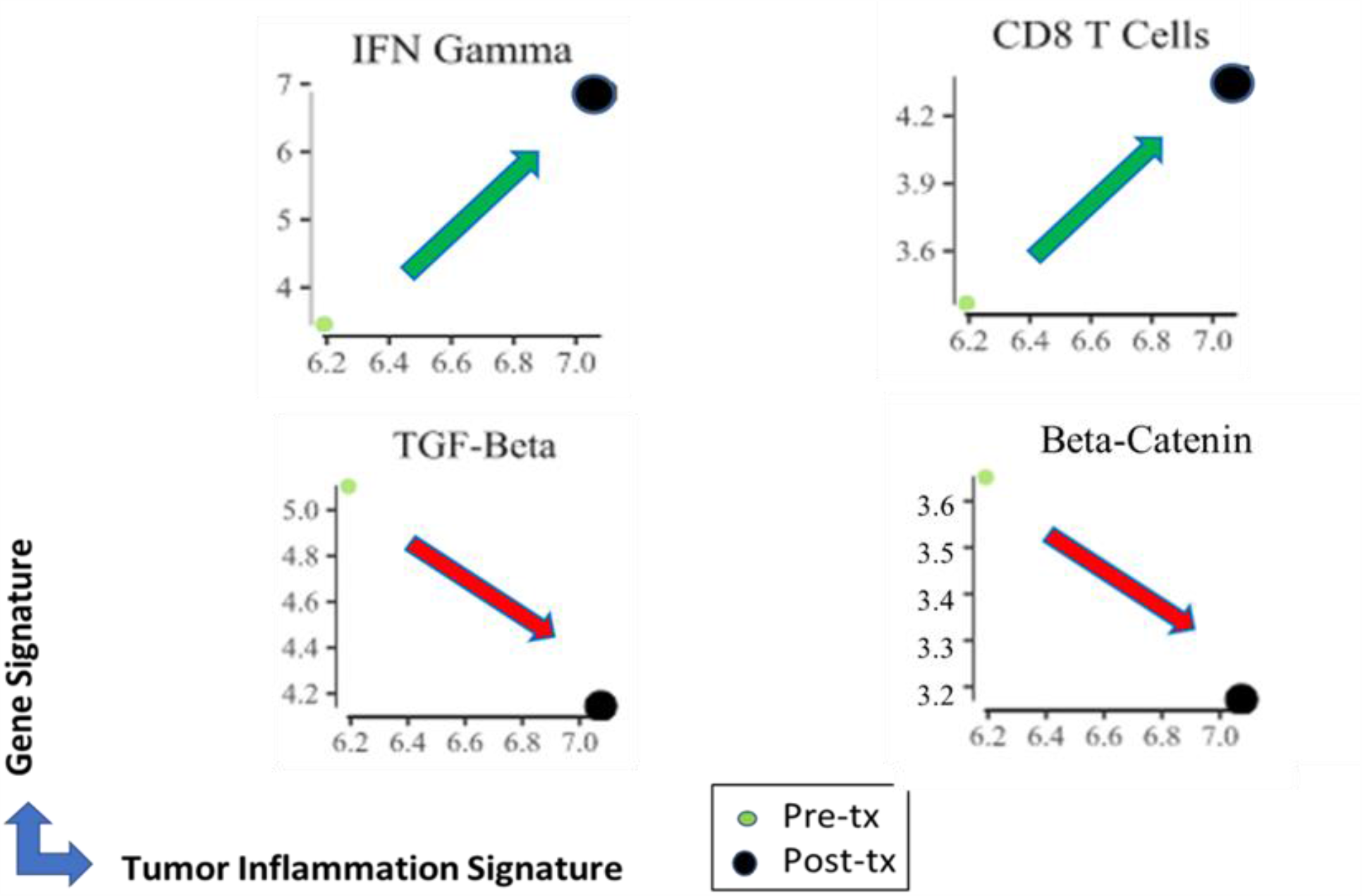
Key changes in pre and post treatment RNA gene expression transcriptome analyses by nanostring IO 360 panel. Ad-p53 treatment increased the Tumor Inflammation Signature and Type I Interferon signaling, decreased TGF-beta and beta-catenin signaling resulting in an increased CD8+ T cell signature which is associated with increased clinical responses to immune checkpoint inhibitor therapy.

Following the documentation of an increased CD8+ T cell response by these nanostring results, the patient was treated with external beam radiotherapy. Owing to the patient’s fragile condition, radiation therapy was initiated at the reduced dose of 30Gy which is approximately 50% less than the standard dose of 70Gy. With respect to possible abscopal effects, remarkably, the patient had a dramatic response to radiation with regression of all tumors including those that had not received full Ad-p53 treatment, full radiation therapy or were initially non-responsive to anti-PD-1 treatment.

## Discussion

Our findings have important implications for future p53 targeted cancer treatments and identify fundamental principles to guide Ad-p53 gene therapy immune oncology applications and provide a hypothesis regarding potential mechanisms contributing to radiation abscopal responses.

With respect to principles guiding p53 targeted therapies, our meta-analysis corroborated previous Ad-p53 monotherapy clinical trials results that identified p53 biomarkers predictive of therapeutic efficacy defined by wild type p53 gene sequence, less than 20% p53 positive tumor cells by immunohistochemistry (IHC), or p53 gene mutations that will not inhibit normal p53 function such as gene deletions, truncations, or frame-shift mutations that have non-functional p53 tetramerization domains (7). In our meta-analysis, all target tumor responses by RECIST 1.1 criteria were observed in patients with favorable p53 biomarker profiles. These favorable p53 biomarker profiles exclude patients with high level expression of dominant negative mutated p53 with intact tetramerization domains which form inactive tetramers that inhibit the activity of wild-type p53 (Nemunaitis 2009). Approximately 75% of cancer patients have favorable p53 biomarker profiles although this percentage may vary between different histological types of cancer(8).

The meta-analysis importantly discovered that Ad-p53 dose based upon the injected tumor volume had a critical effect on tumor responses. While the total Ad-p53 dose administered to each patient was uniform in these previous Ad-p53 monotherapy studies, the amount of Ad-p53 delivered to individual tumor sites was subject to considerably variability. In general, the total dose of 2 x 10^12^ viral particles per patient was divided between the patients’ tumors at the investigators’ discretion which lead to different tumors receiving different Ad-p53 doses. In this meta-analysis, we evaluated the dose of Ad-p53 administered per injected tumor volume using a generally accepted formulae for tumor volume based upon the lesion’s bi-dimensional diameters. Tumor responses were critically dependent upon the amount of the Ad-p53 dose in the injected tumor volume. All Ad-p53 treated target lesion responders by RECIST 1.1 criteria had received Ad-p53 doses greater than 7 × 10^10^ viral particles/cm^3^ tumor and there was a statistically significant difference in tumor responses between patients treated with greater than 7 × 10^10^ viral particles/cm^3^ compared to patients treated at lower Ad-p53 doses.

Hence, previous Ad-p53 clinical trials were negatively impacted by both the inclusion of patients with unfavorable p53 biomarker profiles and by the under dosing of Ad-p53 administration at levels below 7 × 10^10^ viral particles/cm^3^. Of the 70 patients with available biomarker data for the recurrent HNSCC meta-analysis, more than half had either unfavorable biomarker profiles or received suboptimal Ad-p53 dose levels.

Recurrent HNSCC patients have nearly a universal need for additional effective treatments. Recently, the Food and Drug Administration granted approvals in recurrent HNSCC to the anti– PD-1 therapies pembrolizumab and nivolumab. However, these novel anti–PD-1 treatments resulted in only a 13.3% to 16.6% response rate, with a median survival of only 7.5 months(10). Hence, recurrent HNSCC patients have urgent unmet medical needs as current treatments are not curative and typically result in short duration improvements in survival.

In this regard, pre-clinical data has demonstrated the ability of Ad-p53 to reverse resistance to anti-PD-1/anti-PD-L1 therapy in cancer models (13) and these findings led to the initiation of a Phase 2 clinical trial of combined Ad-p53 and anti-PD-1 therapy in recurrent HNSCC and other cancers (https://clinicaltrials.gov/ct2/show/NCT03544723). To identify potential mechanisms of Ad-p53 action contributing to enhanced anti-tumor immunity, pre and post treatment biopsies were evaluated by nanostring gene expression assays. The preliminary results report here indicate that Ad-p53 treatment increased Type I interferon signaling with decreased expression of immunosuppressive oncogenes beta-catenin and TGF-beta leading to increased immune inflammation and CD8 positive T cell signatures. These results support the proposed mechanisms of p53 in reversing immune checkpoint inhibitor resistance shown in Figure 4. These findings will need to be corroborated in additional patients.

**Figure 4.**
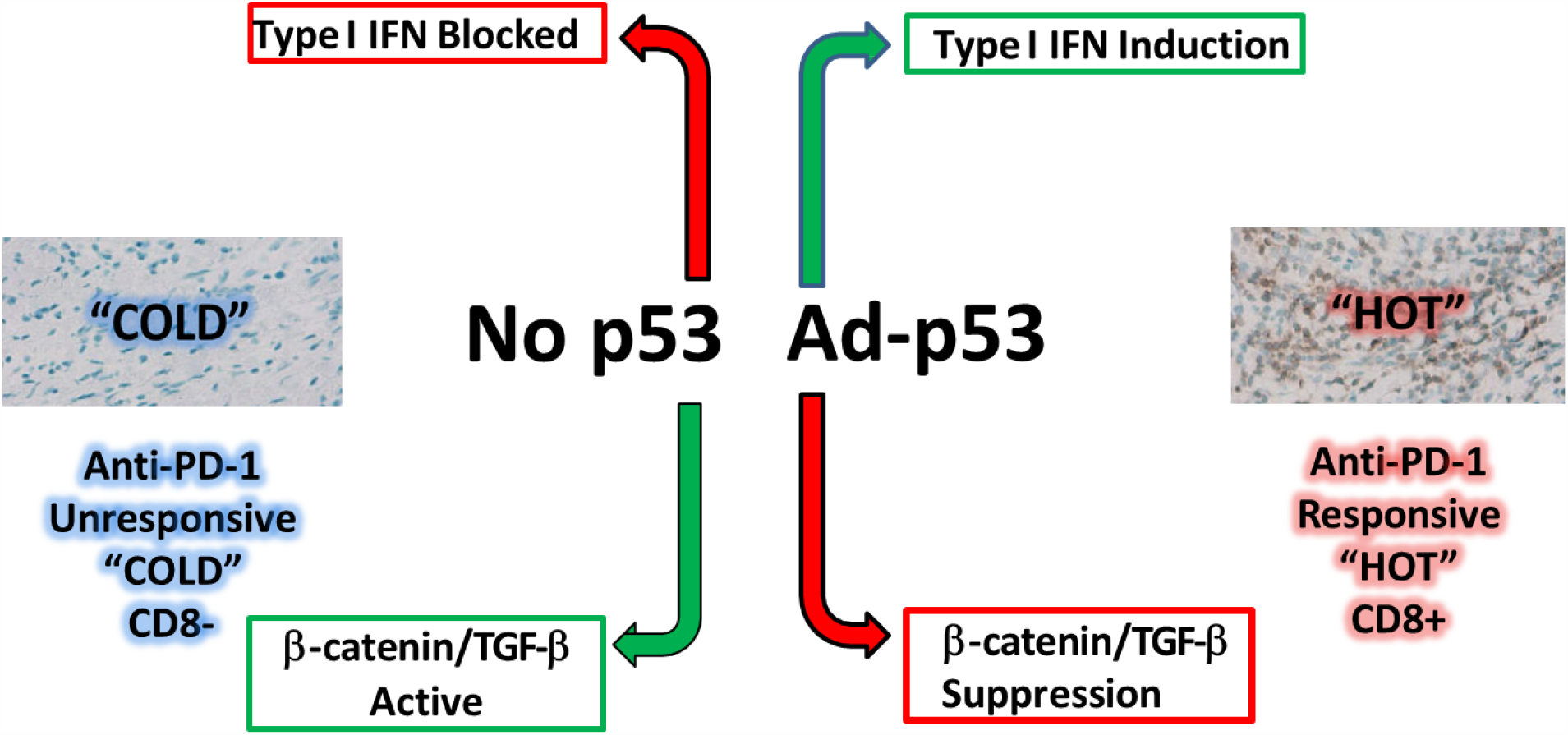
Proposed mechanisms of p53 tumor suppressor immune gene therapy mediated by increased Type I interferon signaling with decreased expression of immunosuppressive oncogenes beta-catenin and TGF-beta leading to increased immune inflammation signatures and CD8 positive T cells.

Our findings also provide a hypothesis regarding potential mechanisms contributing to radiation abscopal responses. The abscopal effect is a very rare and poorly understood phenomenon in the treatment of metastatic cancer where localized treatment of a tumor causes regression of the treated tumor and additional tumors outside the scope of the localized treatment. This phenomenon was first defined in 1953 for radiation therapy by the physician R.H. Mole who proposed the term “abscopal” (‘ab’ - away from, ‘scopus’ - target) to refer to therapeutic effects at a distance from the treated volume but within the same organism (14). Despite its desirability and recognition for over 60 years, it remains a rare and arbitrary phenomenon in the treatment of metastatic cancer.

It is generally believed that abscopal effects result from radiation inducing anti-tumor immune responses which mediate abscopal activity (15) (16) Recent advances in immune oncology have led to widespread use of immune checkpoint inhibitors for cancer therapy. In view of the ability of immune checkpoint inhibitors to amplify anti-tumor immune responses and the presumed relationships between radiation, immune response induction and abscopal effects, it was expected that combination treatments of radiation and immune checkpoint inhibitors would enhance abscopal effects and lead to improved tumor responses for the combined treatment. However, initial reports of combination radiation and immune checkpoint inhibitor treatments have been disappointing and have not increased the therapeutic efficacy above what would be expected with immune checkpoint inhibitor therapy alone. As diagrammed in Figure 5, our results support the hypothesis that abscopal effects are actually memory CD8+ T cell responses that first require inducing or enhancing a CD8+ T cell immune response before administering the localized therapy which triggers the release of antigens leading to activation and expansion of a memory T cell response with abscopal effects. This hypothesis will require validation in future investigations. Previous studies have demonstrated the presence of resident memory T cells in some cancer patients(17).

**Figure 5.**
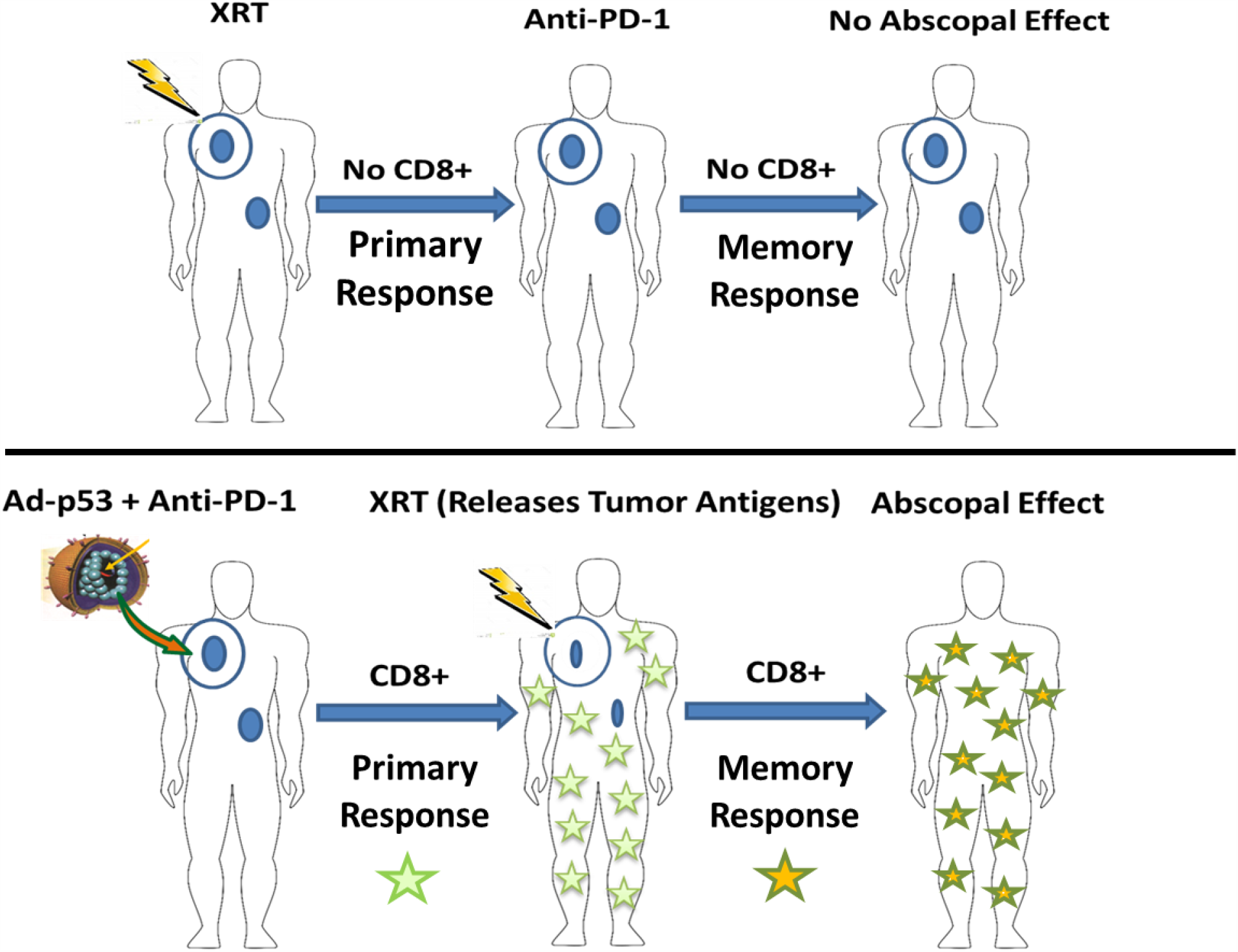
Hypothesis of the abscopal effect as a secondary, memory CD8+ T cell response triggered by antigen release of localized therapy in patients with prior induction of primary anti-tumor CD8+ T cell response.

In summary, our findings have important implications for future p53 targeted cancer treatments and identify fundamental principles to guide Ad-p53 gene therapy immune oncology applications and hypotheses regarding abscopal response mechanisms. We importantly discovered that previous Ad-p53 clinical trials were negatively impacted by both the inclusion of patients with unfavorable p53 biomarker profiles and by the under dosing of Ad-p53 administration levels. Future clinical trials of Ad-p53 should have inclusion criteria for patients with favorable p53 biomarker profiles and Ad-p53 dosing above 7 × 10^10^ viral particles/cm^3^ of injected tumor volume. Our preliminary gene expression profiling results identified p53 mechanisms of action that increased Type I interferon responses and decreased immune suppressive TGF-beta and beta-catenin oncogenes supporting further evaluation of Ad-p53 in combination with immune checkpoint inhibitors to potentially reverse immune therapy resistance in patients with aberrant p53 function. Our results also generated the hypothesis that abscopal effects of localized treatments may reflect systemic memory CD8+ T cell immune responses that first require the effective induction of primary CD8+ T cell anti-tumor immunity with subsequent local treatment releasing antigens that trigger a systemic memory T cell response and abscopal effects. All of our findings will require validation in future randomized, controlled clinical trials.

## Data Availability

The data supporting the conclusions of this article are either incorporated in the manuscript, its supplemental section or available by the corresponding author upon reasonable request.

## Acknowledgements

The authors wish to acknowledge the assistance and support of Nicholas Puro, Casey McCandless, Philip Seaton and William B. Wells. Without their efforts, this work would not have been completed.

## Author Contributors

RES, SC, KBM, and DW designed the research studies. JN conducted the experiments. RES, SC, KBM, MT, BS and DW analyzed the data. RES, SC, KBM, DW and JN wrote the manuscript. All authors reviewed the results and approved the final version of the manuscript.

## Ethics Statement

The studies involving human participants were reviewed and approved by the ethics committee of the University of Toledo and other clinical trial centers where the patients were treated. The patients/participants provided their written informed consent to participate in the clinical trials analyzed in this study.

